# Impairment of endothelial MerTK accelerates atherosclerosis development

**DOI:** 10.1101/2025.04.14.25325845

**Authors:** Shijie Liu, Jingke Yao, Hongye Huang, Jinzi Wu, Oishani Banerjee, Bingzhong Xue, Hang Shi, Zufeng Ding

## Abstract

**Objective:** Atherosclerosis is a chronic inflammatory disease primarily affecting large arteries and is the leading cause of cardiovascular disease. MER proto-oncogene tyrosine kinase (MerTK) plays a key role in regulating efferocytosis, a process for the clearance of apoptotic cells. This study investigates the specific contribution of endothelial MerTK to atherosclerosis development.

**Approach and Results:** Big data analytics, human microarray analyses, proteomics, and a unique mouse model with MerTK deficiency in endothelial cells (*MerTK^flox/flox^Tie2^Cre^*) were utilized to elucidate the role of endothelial MerTK in atherosclerosis development. Our big data analytics, encompassing approximately 98881 cross analyses including 234 analyses for atherosclerosis in the aortic arch, along with human microarray data, reveal that inflammatory responses play a predominant role in atherosclerosis. In vivo, *MerTK^flox/flox^Tie2^Cre^* mice and the littermate control *MerTK^flox/flox^* mice were used to establish an early stage of atherosclerosis model through a high-fat diet combined with AAV8-PCSK9 treatment. Consistent with big data analytics and human microarray analyses, our proteomics data showed that *MerTK^flox/flox^Tie2^Cre^*mice demonstrated significantly enhanced proinflammatory signaling, mitochondrial dysfunction, and activated mitogen-activated protein kinase (MAPK) pathway compared to that of *MerTK^flox/flox^* mice. Endothelial MerTK deficiency induces endothelial dysfunction (enhanced endothelial inflammation, mitochondrial dysfunction, and activation of NADPH oxidases and MAPK signaling pathways) and subsequently causes smooth muscle cell (SMC) phenotypic alterations, ultimately promoting atherosclerosis development.

**Conclusions:** Our findings provide strong evidence that endothelial MerTK impairment serves as a novel mechanism in promoting atherosclerosis development.

## Introduction

Atherosclerosis is an immunoinflammatory disease characterized by lipid accumulation within medium-sized and large arterial walls.^1^ Atherosclerosis is a leading cause of cardiovascular diseases including myocardial infarction, stroke and other plaque-related serious conditions.^1^ Recent studies showed that inflammation responses, dysfunctions in endothelial cells (ECs) and smooth muscle cells (SMCs), and macrophages infiltration play critical roles in the development of atherosclerosis.^1–3^ The vascular endothelium is a monolayer of ECs that provides a permeable barrier to protect vascular vessels from the harmful substances in the blood.^4^ ECs have many important physiological functions, such as maintaining blood fluidity, regulating inflammatory responses and producing vasoactive molecules.^4^ Endothelial dysfunction, characterized by increased oxidative stress, heightened endothelial inflammation, decreased expression of endothelial nitric oxide synthase (eNOS), and reduced nitric oxide (NO) bioavailability, contributes to atherosclerosis initiation and progression.^4,5^ In addition, aortic SMCs display remarkable phenotypic plasticity, transitioning from contractile phenotypes to inflammatory and synthetic phenotypes in response to environmental changes.^6,7^ Endothelial dysfunction and SMC phenotypic alterations are the hallmark features of atherosclerosis.^4–7^

Atherosclerosis can be directly triggered by several mediators, such as accumulation of apoptotic cells, chronic and non-resolving inflammation, and plaque progression, all of which are involved in efferocytosis.^8,9^ Efferocytosis is a precise process in which phagocytes are recruited to the apoptotic tissue, where they recognize, engulf, and clear dying cells in an immunologically silent manner. ^8,9^ Efficient efferocytosis promotes the clearance of apoptotic cells before necrotic core formation and prevents atherosclerosis development; while impaired efferocytosis induces the accumulation of apoptotic cells that in turn accelerates atherosclerotic plaque formation.^8,9^ Mer Tyrosine Kinase (MerTK), a main receptor mediating efferocytosis, is expressed by both professional phagocytes such as macrophages and non-professional phagocytes such as ECs.^8–11^ Our recent studies demonstrated that primary aortic ECs have a high ability to perform efferocytosis via MerTK and play an important role in aortic aneurysms and dissections, a disease that are closely associated with atherosclerosis.^12–15^ Importantly, ECs interact with the neighboring SMCs in vascular systems, potentially influencing SMC functions.^4–7^ However, the specific role of endothelial MerTK in the pathogenesis of atherosclerosis is unknown.

In this study, we utilized integrative big data analytics and microarray analyses of human atherosclerotic tissues, along with our proteomic profiling and an in vivo endothelial specific MerTK-deficient mouse model (*MerTK^flox/flox^Tie2^Cre^*mice), to investigate the role of endothelial MerTK in atherosclerosis. Our data provides compelling evidence that endothelial MerTK impairment significantly accelerates atherosclerosis development, suggesting therapeutic restoration of endothelial MerTK function as a promising strategy to prevent atherosclerosis.

## Methods

### Animals

The *MerTK^flox/flox^* mice and *Tie2-Cre* mice on the C57BL/6J background were generated by Cyagen US (Santa Clara, CA) and housed in the Division of Laboratory Animal Medicine at our institution. All experimental procedures were performed in accordance with protocols approved by the Institutional Animal Care and Use Committee and conformed to the Guidelines for the Care and Use of Laboratory Animals published by the US National Institutes of Health. The mice were maintained under a 12-hour dark cycle at a controlled temperature of 21 ± 1 °C, with ad libitum access to water and a standard laboratory diet. *MerTK^flox/flox^Tie2^Cre^* mice with MerTK conditional knockout in ECs were generated by crossing *MerTK^flox/flox^* mice with *Tie2-Cre* mice (Cyagen US, Santa Clara, CA). The 3-month-old male *MerTK^flox/flox^Tie2^Cre^*mice (n=8) and their littermate control *MerTK^flox/flox^* mice (n=8) were injected with a single dose of AAV8-PCSK9 particles (1×10^11^) via the tail vein, followed by feeding with a high fat diet (HFD, TD.88137, Envigo) for two months to establish an early stage of atherosclerosis model. After mice were euthanized by CO_2_ asphyxiation, the aortic arch tissues were carefully dissected from surrounding tissue, fixed with 10% neutral buffered formalin solution (Sigma, HT501128) and embedded in paraffin for immunohistochemical evaluation or stored in -80 °C for proteomics analysis.

### Genotyping

Genomic DNA was extracted from the tails of the mice using Platinum™ Direct PCR Universal Master Mix (Invitrogen) according to the manufacturer’s instructions. PCR was performed with a 20 μL reaction volume containing 10 µL 2x Platinum™ Direct PCR Universal Master Mix, 1-2 µL sample supernatant, 18–19 µL nuclease-free water, and 0.2 µM primers for *MerTK^flox/flox^*gene of F1: 5’-TTACCTCATGGTATCTGCTGGCTA-3’; R1: 5’-ACCACTTTCTCTT TGGTTGGAGT-3’ and *Tie2-Cre* gene of F1: 5’-CCCTGTGCTCAGACAGAAATGAGA-3’; R1: 5’-CGCATAACCAGTGAAACAGCATTGC-3’. The products were separated by 1.5 % agarose gel electrophoresis and visualized with SYBR™ Safe DNA Gel Stain (Invitrogen) by Syngene™ NuGenius Gel Immaging System.

### Proteomics measurements in the aortic arch samples

Proteomics in the aortic arch samples (n=3 per group) were performed at the Georgia Tech Systems Mass Spectrometry Core. Following data acquisition, proteomic data were analyzed using an empirically corrected library and a quantitative analysis to obtain a comprehensive proteomic profile. Protein identification and quantification were performed using EncyclopeDIA^16^, with results visualized with Scaffold DIA applying 1% false discovery thresholds at both the protein and peptide levels. Protein MS2 exclusive intensity values underwent quality assessment using ProteiNorm.^17^ The data were normalized by cyclic loss to perform statistical analysis using linear models for microarray data (limma) with empirical Bayes (eBayes) smoothing to the standard errors.^25^ Proteins with an FDR adjusted p-value < 0.05 and a fold change > 2 were considered significant. The proteomics data were analyzed by Qiagen IPA Software. All the IPA analyses for proteomics are available with detailed information in the Supplemental Data.

### Big data analytics and human microarray in atherosclerosis

The big data analytics were derived from QIAGEN Ingenuity Pathway Analysis (IPA) and QIAGEN OmicSoft Land Explorer (OLE), which incorporate data from GEO (Gene Expression Omnibus), SRA (Sequence Read Archive), and ArrayExpress. For big data analytics, we set ‘atherosclerosis’ as the key word in IPA Diseases and Functions as well as in IPA Pathways and Lists. For human microarray data evaluation, we derived the data of GSE28829 with seventeen atherosclerotic tissue samples from carotid arteries (n = 9 in early and n = 8 in advanced lesions).^18^ The IPA comparisons were performed between late-stage and early-stage atherosclerosis. For IPA Analysis Match between our proteomics and public shared projects in atherosclerosis, we filtered the data with the key words of ‘atherosclerosis’ in case.diseasestate and ‘aorta’ in case.tissue. We identified 16 projects in atherosclerosis from different mouse models: 3- atherosclerosis [aorta] NA CMP_ckHSOYUqU4ic; 4- atherosclerosis [aorta] NA CMP_vOcWhcD4rOLk; 7- atherosclerosis [aorta] NA CMP_Rvm3t858gPeo; 8- atherosclerosis [aorta] NA CMP_I3FNY2ilGPYM; 21- atherosclerosis [aorta] NA CMP_CTtNehHyXKgd; 20- atherosclerosis [aorta] NA CMP_GJ6Fs9Oo9osL; 2- atherosclerosis [aorta] NA CMP_MToofiTDMtxi; 2- atherosclerosis [aorta] NA CMP_7ym4dlpKMoKs; 6- atherosclerosis [aorta] NA CMP_kplAZ21kAllP; 24- atherosclerosis [aorta] NA CMP_I4YzR4eT1K2r; 5- atherosclerosis [aorta] NA CMP_wI600H3ALvA7; 2- atherosclerosis [aorta] NA CMP_OEkcJYZWVInc; 19- atherosclerosis [aorta] NA CMP_ujRjt7XjNWqY; 1- atherosclerosis [aorta] NA CMP_QNA489a6316v; 22- atherosclerosis [aorta] NA CMP_j13s8fMql4l4; 23- atherosclerosis [aorta] NA CMP_G2C5o1Lau1XD. All the IPA analyses are available with detailed information in the Supplemental Data.

Based on QIAGEN IPA citation guidelines, a data set containing gene identifiers and corresponding data measurement values was uploaded into the application. Each identifier was mapped to its corresponding entity in QIAGEN’s Knowledge Base. Network Eligible molecules were overlaid onto a global molecular network developed from information contained in the QIAGEN Knowledge Base. Networks of Network Eligible Molecules were then algorithmically generated based on their connectivity. The Diseases & Functions Analysis identified the biological functions and/or diseases that were most significant from the data set. Molecules from the dataset that were associated with biological functions and/or diseases in the QIAGEN Knowledge Base were considered for the analysis. A right-tailed Fisher’s Exact Test was used to calculate a p-value determining the probability that each biological function and/or disease assigned to that data set is due to chance alone. A z-score was calculated to indicate the likelihood of an increase or decrease of that disease or function. There are about 1500 diseases, phenotypes, and function pathways created by Machine Learning (ML) in the QIAGEN Knowledge Base. These ML Disease Pathways show key molecules that impact a single disease and its associated phenotypes, which may represent novel participants in the disease or its etiology.

### Immunofluorescent staining

Immunofluorescent staining was performed according to the protocol of ‘Immunofluorescent Staining of Paraffin-Embedded Tissue (Novus Biologicals)’. The information of antibodies is shown as follows: IL-1β, p22^phox^, p38 and α-SMA were from Cell Signaling; MCP-1 and TNF-α were from Invitrogen; ApoE was from Proteintech; p47^phox^ and gp91^phox^ were from SantaCruz Biotechnology; IFN-γ was from BiossUSA; and JNK was from Proteintech.

### Statistical analysis

An unpaired Student’s t-test was used to determine statistical significance between two groups. Dunnett’s one-way ANOVA was used for multiple comparisons between disease types and normal control. Data was analyzed with GraphPad Prism 9.4.1 and summarized as the mean ± SD. *P* < 0.05 was considered statistically significant.

## Results

### Big data analytics and microarray analysis of human atherosclerotic tissues

Atherosclerosis is characterized by chronic inflammation and lipids accumulation within large arteries.^1–3^ While extensively studied in various animal models (rodents, pigs, rabbits) and human tissue samples, the comprehensive signaling pathways remain complex. Using Qiagen IPA Machine Learning Disease Pathways, we analyzed atherosclerosis-associated data from 98,881 cross analyses, including 234 analyses specifically focused on the aortic arch (**Figure 1A**-**B**). Machine Learning identified approximately∼1500 disease-associated pathways, highlighting significant activation of inflammatory signaling (e.g., IL-1β, CCL2, CXCL2, CCR2, ACKR1) and angiotensinogen (AGT), along with suppression of lipid-related pathways involving ApoE and LDLR. (**Figure 1C**). To further reveal the common signaling that are involved in atherosclerosis, we next performed microarray reanalysis of human atherosclerotic tissues. Our data indicates significant changes of upstream pathways including both top 50 inhibited signaling (e.g., ApoE, SOCS1, and IL1RN) and top 50 activated signaling (TNF, IFN-γ, and IL1 family) (**Figure 1D**-**E**). Consistent with these findings, the overall signaling of microarray analysis of human atherosclerotic tissues revealed activated inflammatory markers such as IL-1β, CSF2, IFN-γ, TLR2/7/9, TNF, and TNFSF13B (**Figure 1F**). MerTK is the main receptor for efferocytosis and plays an important role in cardiovascular disease (CVD) including atherosclerosis, myocardial infarction and hypertension.^8,9^ To further reveal the key role of MerTK in cardiovascular diseases, we performed the second big data analytics focusing on aortic tissues of human cardiovascular diseases based on RNA-seq data from IPA database with a total of 427 samples. Our data showed that, compared to normal control group, MerTK is higher expressed in embryotic aorta while it is significantly inhibited in cardiovascular diseases, particularly respiratory tract disease (RTD)-related cardiovascular diseases (**Figure 1G**). MerTK is highly expressed in professional phagocytes such as macrophages and other monocytes. However, the BioGPS data showed that MerTK mRNA expression is also highly expressed in vascular ECs compared to bone marrow (macrophages origination) and monocytes (**Figure 1H**). In summary, our big data analytics and microarray reanalysis using human atherosclerotic tissues confirmed that activated inflammation responses and lipids accumulation are the main characteristics of atherosclerosis. The RNA-seq big data analytics and BioGPS data emphasized endothelial MerTK may play a novel role in atherosclerosis. Importantly, recent studies from us and other research groups have confirmed that aortic ECs can highly express MerTK and play a key role in aortic aneurysms and dissections, a condition closely associated with atherosclerosis. This promotes us to focus on endothelial MerTK and investigate its role in atherosclerosis development.

**Figure 1.**
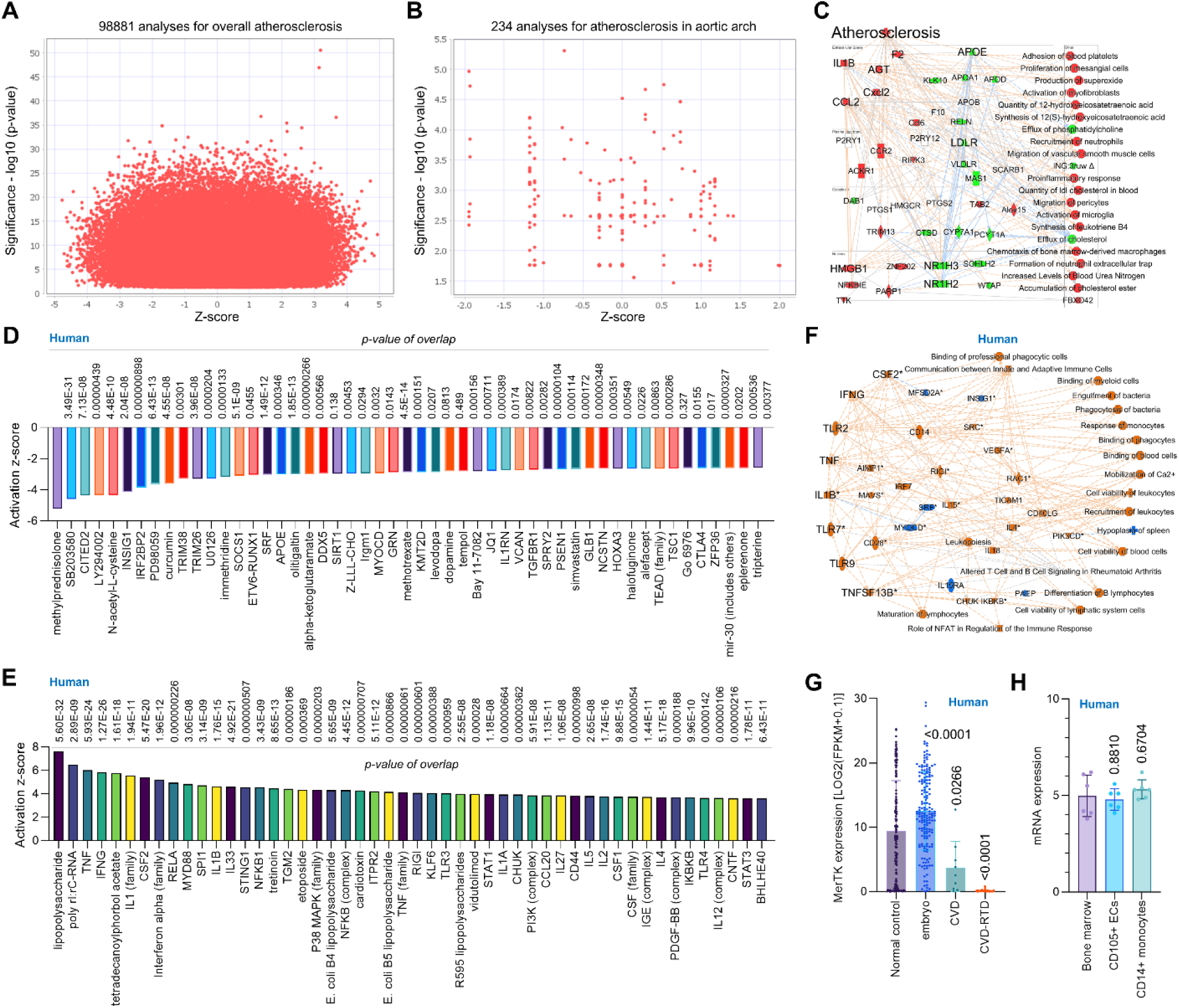
Big data analytics and human microarray in atherosclerosis. *Big data analytics.* (**A**-**C**) Big data analytics for atherosclerosis with 98881cross analyses for overall signaling and 234 cross analyses in aortic arch based on up-to-date RNA-seq data from humans, mouse and rat. In IPA of Pathways and Lists, atherosclerosis was set as the keywords. ***Microarray in human atherosclerosis.*** (**D**-**E**) The top 50 downregulated or upregulated upstream regulators based on activation of z-score. (**F**) Graphical summary of human microarray data (orange: upregulated; blue: downregulated). QIAGEN Ingenuity Pathway Analysis (IPA: 1-atherosclerosis [carotid atherosclerotic plaque] NA CMP_2gGgljQ5SpJAn) and QIAGEN OmicSoft Land Explorer (OLE) were used to analyze microarray data in carotid atherosclerotic plaque from human patients. ***RNA-seq big data analytics in human***. (**G**) MerTK expression in human diseases specifically in related aortic tissues (n=427 in total), including normal control, embryo, cardiovascular disease (CVD) and respiratory tract disease (RTD)-related cardiovascular disease. MerTK expression was based on RNA-seq or scRNA-seq and was quantified by Log2 (FPKM + 0.1). Original data of RNA-seq or scRNA-seq for MerTK expression were downloaded from QIAGEN OmicSoft Land Explorer. ***BioGPS***. (**H**) MerTK mRNA expression in human cells derived from BioGPS (http://biogps.org). The data were analyzed with GraphPad Prism 9.4.1 and shown as the mean ± SD.

### Endothelial MerTK deficiency promotes atherosclerosis

To explore the role of endothelial MerTK in atherosclerosis and validate the signaling pathways from big data analytics and human microarray analysis, we established an early stage of atherosclerosis model in *MerTK^flox/flox^* mice and *MerTK^flox/flox^Tie2^Cre^* mice. Body weight measurement showed no significant differences between *MerTK^flox/flox^*mice and *MerTK^flox/flox^Tie2^Cre^* mice (**Figure 2A**). However, obvious atherosclerotic plaque formation was found in *MerTK^flox/flox^Tie2^Cre^*mice whereas no obvious plaque formation was detected in the control *MerTK^flox/flox^*mice (**Figure 2B**-**C**). Given the key role of inflammation in atherosclerosis, we performed immunostaining to assess the expression of inflammatory markers IL-1β, MCP-1, NF-κB, TNF-α, IFN-γ. IL-1β, a key pro-inflammatory cytokine implicated in atherosclerosis progression and plaque formation^19^, was widely expressed in the aortic arch of both *MerTK^flox/flox^* mice and *MerTK^flox/flox^Tie2^Cre^*mice but was predominantly localized to the endothelium of *MerTK^flox/flox^Tie2^Cre^*mice (**Figure 2D**). Monocyte chemoattractant protein-1 (MCP-1), a chemokine essential for monocytes/macrophages migration and infiltration^20^, exhibited significantly increased expression in the aortic arch of *MerTK^flox/flox^Tie2^Cre^* compared to *MerTK^flox/flox^* mice (**Figure 2E**). NF-κB, a pivotal regulator of inflammatory responses responsible for inducing proinflammatory gene expression, including IL-1β and MCP-1^21^, was markedly elevated in *MerTK^flox/flox^Tie2^Cre^* mice (**Figure 2G**). Similarly, TNF-α and IFN-γ, central mediators of proinflammatory responses in atherosclerosis^1–3^, were upregulated in the aortic arch of *MerTK^flox/flox^Tie2^Cre^*mice compared to the control *MerTK^flox/flox^* mice (**Figure 3A, 3D**). ApoE, known for its protective role in clearing lipoproteins from circulation and preventing atherosclerosis, was significantly increased in the aortic arch of *MerTK^flox/flox^* mice but markedly reduced in *MerTK^flox/flox^Tie2^Cre^* mice (**Figure 2F**).

**Figure 2.**
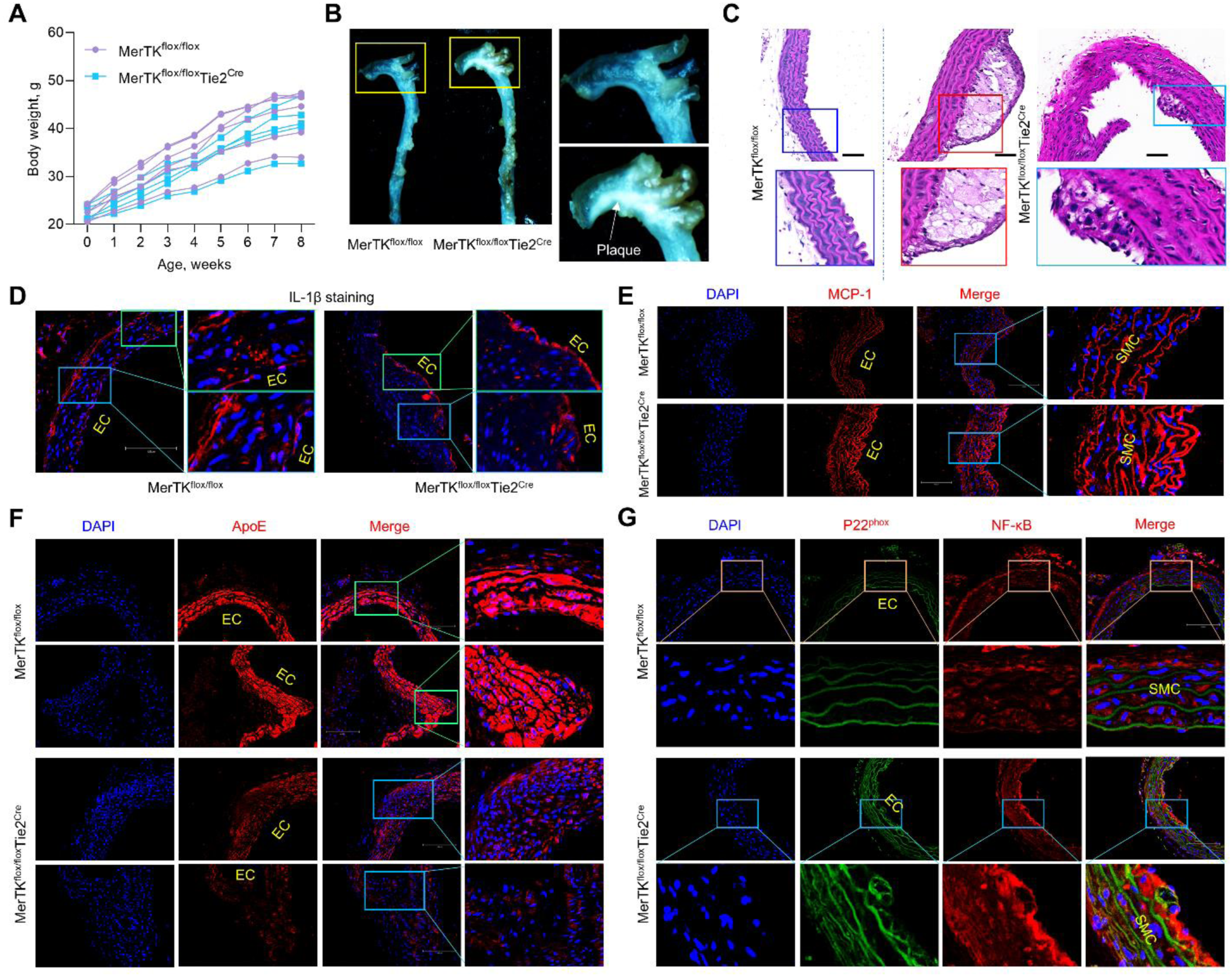
Endothelial MerTK and atherosclerosis. (**A**) Body weight measurement for two months. (**B**-**C**) The visible formation of atherosclerotic plaque and H&E staining in aortic arch. (**D**-**G**) Immunostaining for the expression of IL-1β, MCP-1, ApoE, p22^phox^ and NF-κB in aortic arch from *MerTK^flox/flox^*mice and *MerTK^flox/flox^Tie2^Cre^* mice of atherosclerosis model.

**Figure 3.**
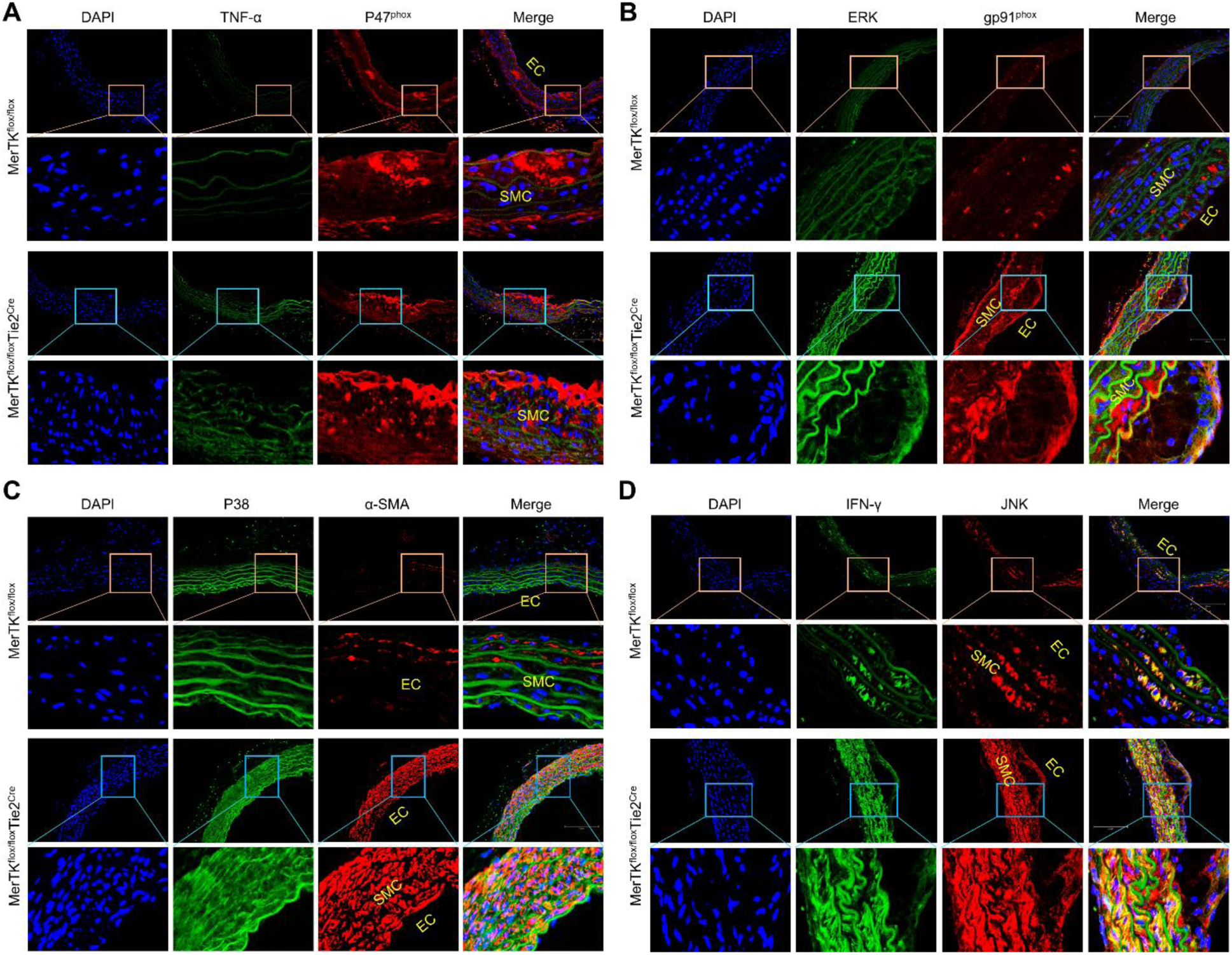
Endothelial MerTK and atherosclerosis. (**A**-**D**) Immunostaining for the expression of inflammation markers (TNF-α and IFN-γ), NADPH oxidase subunits (p47^phox^ and gp91^phox^), and MAPK family (ERK, p38 and JNK) in aortic arch from *MerTK^flox/flox^* mice and *MerTK^flox/flox^Tie2^Cre^* mice of atherosclerosis model.

NADPH oxidases, that generate superoxide or hydrogen peroxide, are a major source of cellular reactive oxygen species (ROS) production.^22^ Increasing evidence indicates that mitochondrial dysfunction promotes NADPH oxidase activation, exacerbating oxidative stress and inflammation, thereby contributing to atherosclerosis progression.^23^ Here, we assessed NADPH oxidase activation as a marker of oxidative stress and mitochondrial dysfunction. Our data revealed significantly increased expression of NADPH oxidase subunits (p22^phox^, p47^phox^, and gp91^phox^) in the aortic arch of *MerTK^flox/flox^Tie2^Cre^* mice compared to *MerTK^flox/flox^* mice (**Figure 2G** and **3A-B**). Interestingly, these NADPH oxidase subunits were predominantly expressed in the ECs of *MerTK^flox/flox^Tie2^Cre^*mice, indicating the importance of endothelial MerTK in regulating NADPH oxidase activation. Similarly, the MAPK signaling pathway, known for its involvement in atherosclerosis, promotes endothelial dysfunction, SMC phenotypic alterations and pro-inflammatory macrophage infiltration. The main MAPK family members include extracellular signal-regulated kinase (ERK), p38 MAPK, and Jun kinase (JNK). Our data revealed significantly higher expression of ERK, p38 MAPK and JNK in the aortic arch of *MerTK^flox/flox^Tie2^Cre^* mice compared to the control *MerTK^flox/flox^* mice (**Figure 3B**-**D**).

During atherosclerotic plaque development, vascular SMCs often undergo phenotypic alterations, adopting proinflammatory characteristics and losing SMC markers.^6,7^ Our data showed that proinflammatory markers, including MCP-1, NF-κB, TNF-α, and IFN-γ, were highly expressed in SMCs from *MerTK^flox/flox^Tie2^Cre^* mice compared to the control *MerTK^flox/flox^* mice (**Figure 2**-**3**). In addition, NADPH oxidase subunits (p22^phox^, p47^phox^, and gp91^phox^) and MAPK family members (ERK, p38 MAPK and JNK), all of which are involved in SMC phenotypic switching in atherosclerosis, were significantly elevated in SMCs of *MerTK^flox/flox^Tie2^Cre^*mice compared to *MerTK^flox/flox^* mice (**Figure 2**-**3**). Α-smooth muscle actin (α-SMA), a well-established marker of contractile SMCs and an indicator of endothelial to mesenchymal transition (EndMT, a hallmark of endothelial dysfunction)^24^ was predominantly expressed in SMCs with almost no detection in the endothelium of the control *MerTK^flox/flox^* mice (**Figure 3C**). However, in the *MerTK^flox/flox^Tie2^Cre^* mice exhibited markedly increased α-SMA expression in both endothelium and SMCs, suggesting that endothelial MerTK deficiency promotes EndMT and SMC phenotypic alterations in atherosclerosis. In summary, our in vivo findings using the unique *MerTK^flox/flox^Tie2^Cre^*atherosclerosis model corroborate the key observations from big data analytics and human microarray analysis. These results underscore the pivotal role of proinflammatory signaling, NADPH oxidases, MAPK signaling pathway, and SMC phenotypic alterations in endothelial MerTK-mediated atherosclerosis.

### Proteomic analysis of endothelial MerTK deficient mice

To further reveal underlying mechanisms, proteomics was employed to investigate protein-protein interactions in endothelial MerTK-mediated atherosclerosis. **Figure 4A** shows the abundance ratios (weights and log2) of *MerTK^flox/flox^Tie2^Cre^*group vs. *MerTK^flox/flox^* group from the original proteomics data analyzed by Proteome Discoverer 3.2 (Thermo Fisher Scientific). To optimize our exploration of the underlying mechanisms of endothelial MerTK in atherosclerosis, we analyzed a total of 1548 proteins (587 down and 961 up) emphasizing 97 proteins with a significant change (24 down and 73 up) using Qiagen IPA software. As shown in **Figure 4B**, there is a clear separation in protein profiles between *MerTK^flox/flox^Tie2^Cre^*mice and *MerTK^flox/flox^* mice. The graphical summary of our proteomics revealed a significant upregulation (e.g., EGF, IGF1, CSF1, HIF1α, MAPK subunits) and downregulation (e.g., LARP1, KLF15, PNPLA2, PPARGC1α) in *MerTK^flox/flox^Tie2^Cre^* mice compared to *MerTK^flox/flox^*mice (**Figure 4C**). Interestingly, our proteomic data highlighted the key role of mitochondrial dysfunction in endothelial MerTK-mediated atherosclerosis (**Figure 4C**). In consistence, our canonical pathway analysis also showed that mitochondrial dysfunction emerged as a critical pathway in endothelial MerTK-mediated atherosclerosis using the big data analytics with 67629 cross analyses (**Figure 4D**). Upstream regulator analysis discovered activated signaling (TGFβ1, HIF1α, VEGF) and inhibited proteins (LARP1, FMR1, PPARGC1α), underscoring mitochondrial dysfunction and MAPK signaling pathways’ importance in endothelial MerTK-mediated atherosclerosis (**Figure 4E**-**4F**). In summary, our proteomic analysis revealed the key roles of MAPK subunits, TGFβ signaling, and mitochondrial dysfunction in endothelial MerTK-mediated atherosclerosis, promoting us to further analyze mitochondrial dysfunction in *MerTK^flox/flox^Tie2^Cre^* mice.

**Figure 4.**
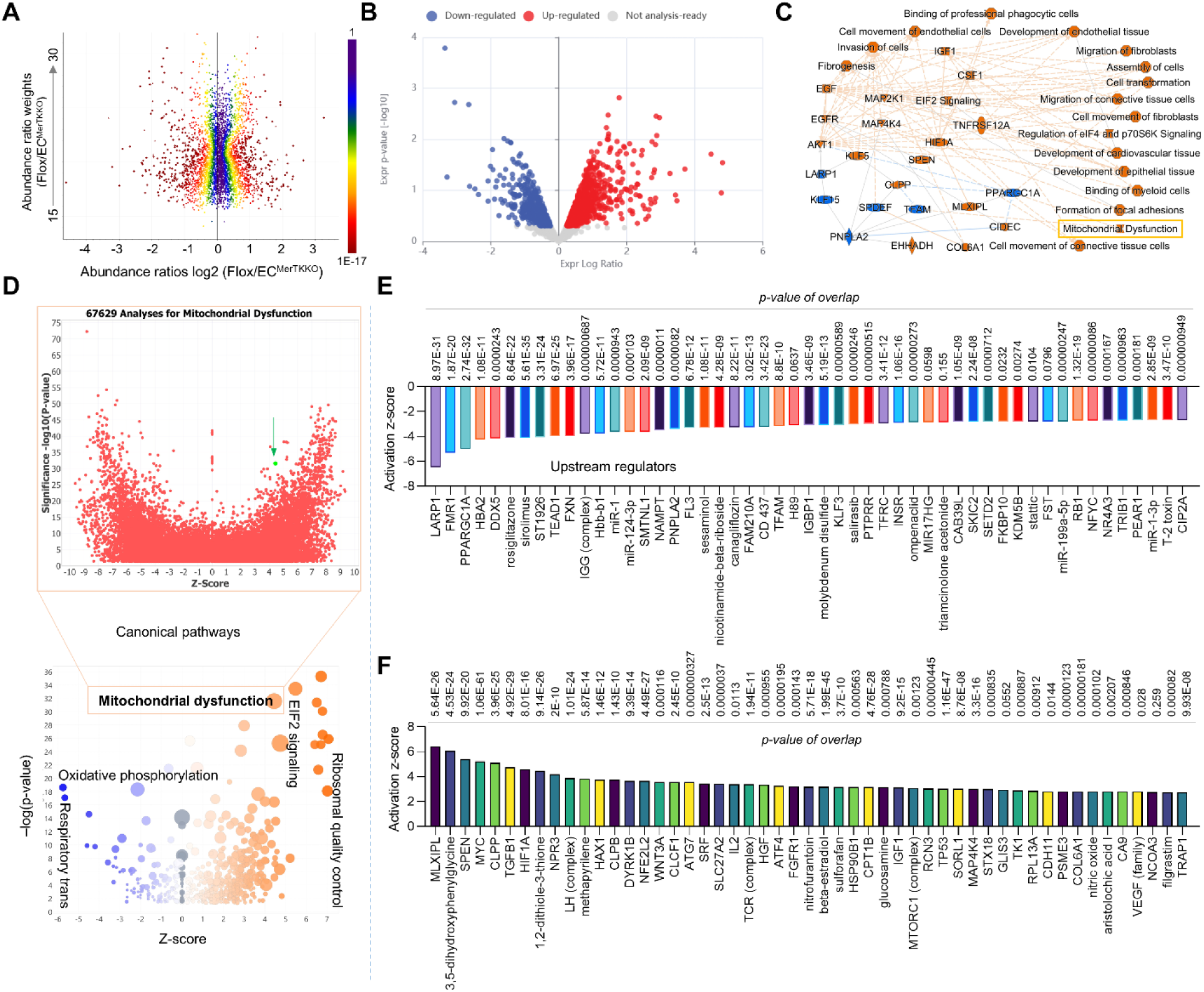
Proteomics in atherosclerotic aortic arch with *MerTK^flox/flox^Tie2^Cre^* vs. *MerTK^flox/flox^*. (**A**) Protein abundance in comparison of *MerTK^flox/flox^* group with *MerTK^flox/flox^Tie2^Cre^* group. (**B**) Volcano plot illustrating differentially expressed proteins in the aortic arch of *MerTK^flox/flox^Tie2^Cre^* vs. *MerTK^flox/flox^*. Relative protein abundance (log2) plotted against significance level (-log10 P-value), showing downregulated (blue), upregulated (red) or non-differentially expressed proteins (gray). (**C**) Graphical summary of proteomics data (orange: upregulated; blue: downregulated). (**D**) The volcano canonical pathways based on activation of z-score (lower panel). Blue: negative value. Orange: positive value. Grey: no activity pattern. Size is based on the number of genes that overlap the pathway. Big data analytics for 67629 cross analyses for mitochondrial dysfunction based on IPA data base (upper panel). (**E**-**F**) The top 50 downregulated or upregulated upstream regulators based on activation of z-score.

### Endothelial MerTK deficiency promotes mitochondrial dysfunction in atherosclerosis

Mitochondrial dysfunction promotes chronic inflammation responses, accumulation of lipids, apoptotic cell buildup, and the plaque necrotic core formation, leading to the initiation and progression of atherosclerosis.^25^ IPA Machine Learning analysis leveraging the QIAGEN Knowledge Base provides an opportunity to uncover novel relationships between our proteomics and disease phenotypes. Here, we specifically assessed mitochondrial function to determine its contribution to endothelial MerTK-mediated atherosclerosis. **Figure 5A** presents Machine Learning-derived disease pathways by comparing *MerTK^flox/flox^Tie2^Cre^*mice and their littermate control *MerTK^flox/flox^* mice. Notably, *MerTK^flox/flox^Tie2^Cre^* mice exhibited significantly increased mitochondrial dysfunction, including mitochondrial DNA-related disorders, mitochondrial diseases, mitochondrial cytopathy and mitochondrial myopathy. **Figure 5B** shows sixteen molecules associated with mitochondrial DNA-related disorders, including inhibited signaling pathways (e.g., SIRT3, LDHB, and SURF1) and activated pathways (NQO1, PHGDH, and ANK3). **Figure 5C**-**5F** illustrates IPA network analysis detailing overall signaling pathways involved in endothelial MerTK-mediated atherosclerosis, highlighting the candidate proteins, the protein-protein interactions, and the signaling pathways such as increased glutamine levels and oxygen consumption. **Figure 5G**-**5H** shows IPA predictions identifying molecules upregulated (e.g., APT1A3, Gsta1, and Calm4) or inhibited (e.g., SDHD, SIRT3, and RHOT1) in *MerTK^flox/flox^Tie2^Cre^*mice compared to the control *MerTK^flox/flox^* mice. In summary, our Machine Learning disease pathway analyses reveal the crucial role of mitochondrial dysfunction in endothelial MerTK-mediated atherosclerosis.

**Figure 5.**
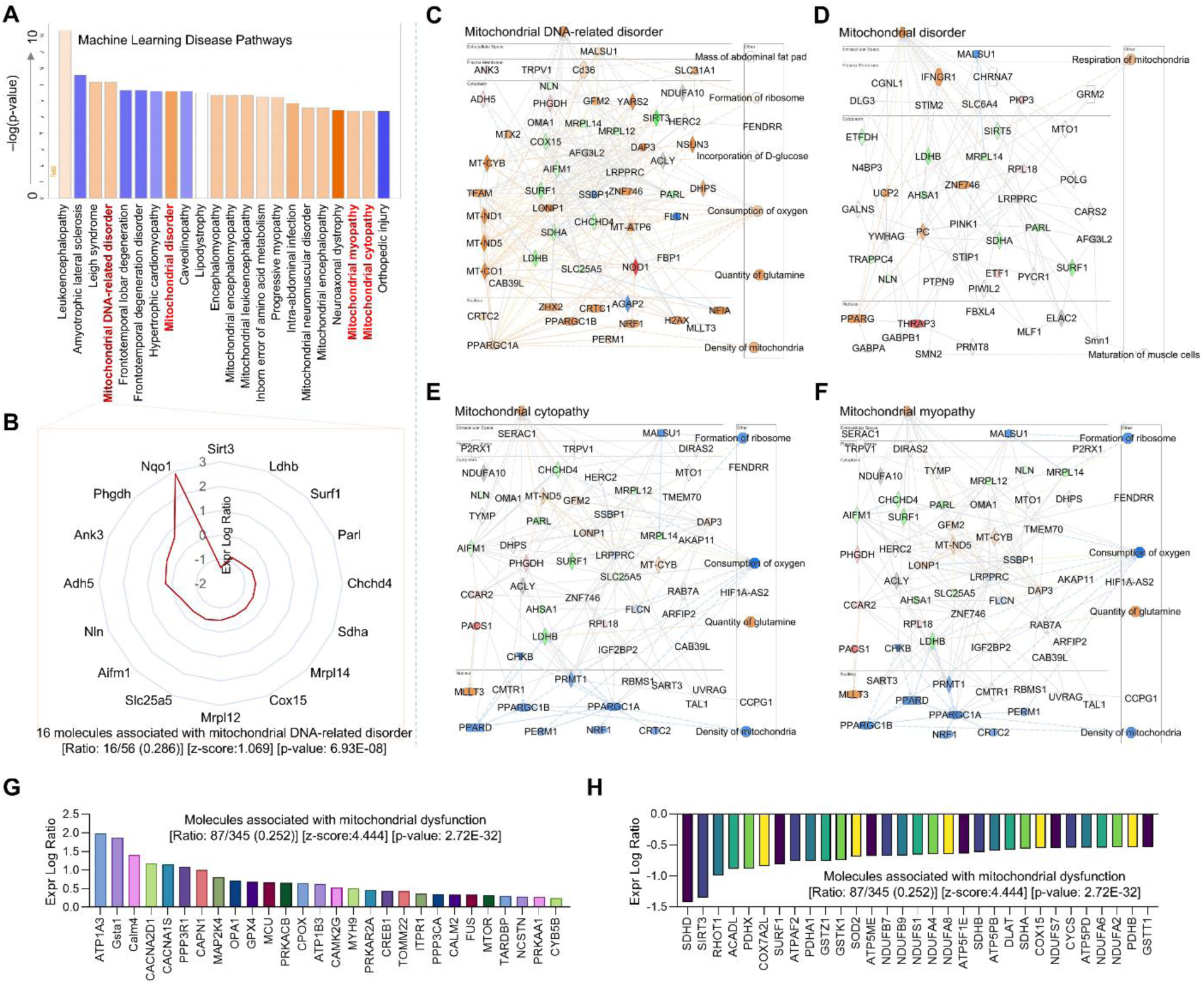
Proteomics focusing on mitochondrial dysfunction in *MerTK^flox/flox^Tie2^Cre^*group compared to *MerTK^flox/flox^* group. (**A**-**B**) Machine learning (ML) disease pathways showing mitochondrial dysfunctions and the protein changes of mitochondrial DNA-related disorder (orange: upregulated; blue: downregulated). (**C**-**F**) Graphical summary for mitochondrial DNA-related disorder, mitochondrial disorder, mitochondrial cytopathy, and mitochondrial myopathy. Red: increased measurement. Green: decreased measurement. Orange: predicted activation. Bule: predicted inhibition. Glos indicates activity when the opposite of measurement. The lines indicate the predicated relationship (orange: leads to activation; blue: leads to inhibition; yellow: findings inconsistent with state of downstream molecule; grey: effect not predicted). (**G**-**H**) Activated or inhibited molecules associated with mitochondrial dysfunction.

### Causal network analysis reveals the importance of MAPK in the endothelial MerTK deficiency-mediated atherosclerosis

The IPA causal network analysis provides a comprehensive approach to identify key master regulators and novel regulatory mechanisms using the Ingenuity Knowledge Base. As described in Qiagen IPA website, the Ingenuity Knowledge Base is a curated repository of biological interactions and functional annotations derived from millions of individually modeled relationships among proteins, genes, complexes, cells, tissues, drugs, and diseases. As shown in **Figure 6A**, our causal network identified a variety of upregulated signaling pathways including MAPK family and TGFβ family in *MerTK^flox/flox^Tie2^Cre^*mice. In consistence, the IPA analysis also predicted the networks of MAPK family and TGFβ family (**Figure 6A**), highlighting the interactions between MAPK signaling and TGFβ signaling. Interestingly, our causal network analysis for the top 50 inhibited signaling further showed that endothelial MerTK deficiency effectively blocks the effects of MAPK inhibitors, including SB203580 (a potent inhibitor of p38 MAPK)^26^ (**Figure 6B**). In addition to MAPK signaling and TGFβ signaling, our causal network analysis identified several microRNAs (miR) involved in endothelial MerTK deficiency-mediated atherosclerosis. These include inhibited miRs such as miR-217-5p, miR-451, and miR-3133 and activated miRs such as miR-218, miR-199a-5p, and miR-185 (**Figure 6C**). Of note, many of these miRs regulate MAPK signaling pathway as shown in the representative miR-218 with IPA prediction (**Figure 6C**). Collectively, these findings reinforce the critical roles of MAPK signaling and TGFβ signaling in endothelial MerTK-mediated atherosclerosis.

**Figure 6.**
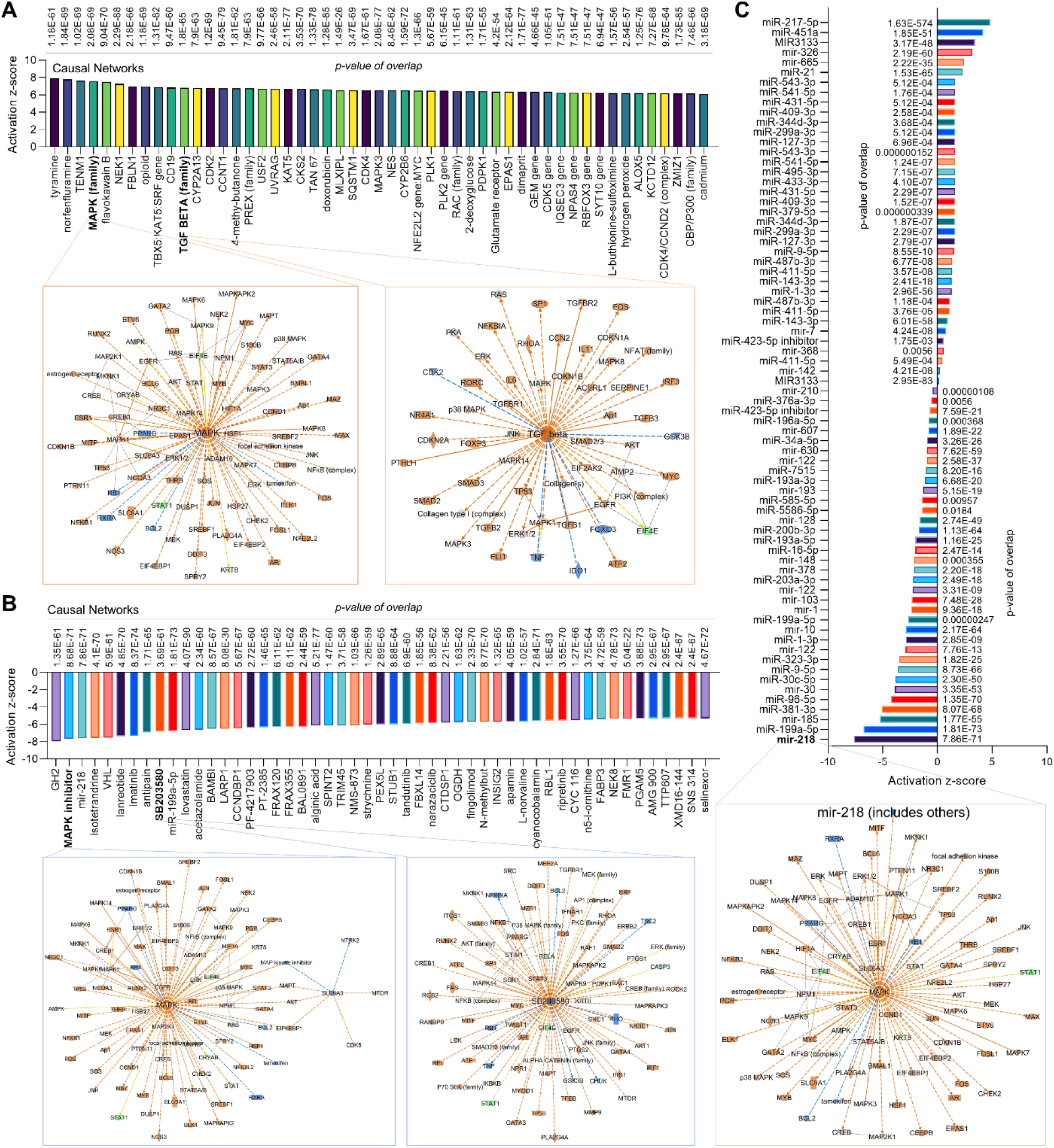
Proteomics causal networks in atherosclerotic aortic arch with *MerTK^flox/flox^Tie2^Cre^* vs. *MerTK^flox/flox^*. (**A**) The top 50 activated upstream regulators and IPA prediction of MAPK family and TGFβ family networks based on activation of z-score. (**B**) The top 50 inhibited upstream regulators and IPA prediction of MAPK inhibitor and SB203580 networks based on activation of z-score. (**C**) The upregulated or downregulated microRNAs in *MerTK^flox/flox^Tie2^Cre^* group compared to *MerTK^flox/flox^*group. Representative IPA prediction micorRNA-218 focusing on MAPK signaling pathway. Upregulated and downregulated proteins are highlighted in red and green, respectively, and the color depth is correlated to the fold change. Orange and blue dashed lines with arrows indicate indirect activation and inhibition, respectively. Yellow and gray dashed lines with arrows depict inconsistent effects and no prediction, respectively.

### Multi-comparisons in different atherosclerosis projects

To further investigate the role of endothelial MerTK in atherosclerosis development, we performed an IPA multi-comparative analyses comparing our *MerTK^flox/flox^Tie2^Cre^*-mediated atherosclerosis models with other atherosclerosis models available from the IPA database. The IPA database encompassed 16 projects, including ApoE^-/-^ versus WT, atherosclerosis versus normal control in young and aged populations, and high-fat diet versus chow diet in WT mice. **Figure 7A** shows top 20 upstream regulator signals across the atherosclerosis models including *MerTK^flox/flox^Tie2^Cre^*(vs. *MerTK^flox/flox^*) and other atherosclerosis projects. We found robust activation of proinflammatory signals commonly associated with atherosclerosis (e.g., TNF, IL1A, and IL1B) and other signaling pathways implicated in atherosclerosis progression (e.g., AGT, IFR2BP2, and PPARGC1A). **Figure 7B** shows the top 20 canonical pathways, highlighting significant enrichment of pathways related to cytokine storm formation, mitochondrial dysfunction, and cell surface interactions at the vascular wall. Interestingly, mitochondrial dysfunction was markedly elevated in *MerTK^flox/flox^Tie2^Cre^*mice compared with other atherosclerosis projects. **Figure 7C** details multi-comparative analysis of toxicity-related functions, indicating that atherosclerosis may cause various toxic effects in other organs, such as liver cell death, left ventricular (LV) enlargement, and renal fibrosis. Compared to other atherosclerosis models, endothelial MerTK deficiency enhanced hepatic inflammation, liver damage, and hepatic steatosis, highlighting a promising area for future research into the role of endothelial MerTK in liver function. **Figure 7D** summarizes IPA multi-comparative analysis for diseases and biological functions between the *MerTK^flox/flox^Tie2^Cre^* model (vs. *MerTK^flox/flox^* mice) and other atherosclerosis models. Consistent with our previous findings on impaired efferocytosis MerTK-deficient aortic ECs, endothelial MerTK deficiency strongly inhibits phagocytotic activities. In summary, these multi-comparative analyses further substantiate the critical role of endothelial MerTK dysfunction in atherosclerosis, with endothelial MerTK-mediated phagocytosis including efferocytosis, emerging as a central pathogenic mechanism.

**Figure 7.**
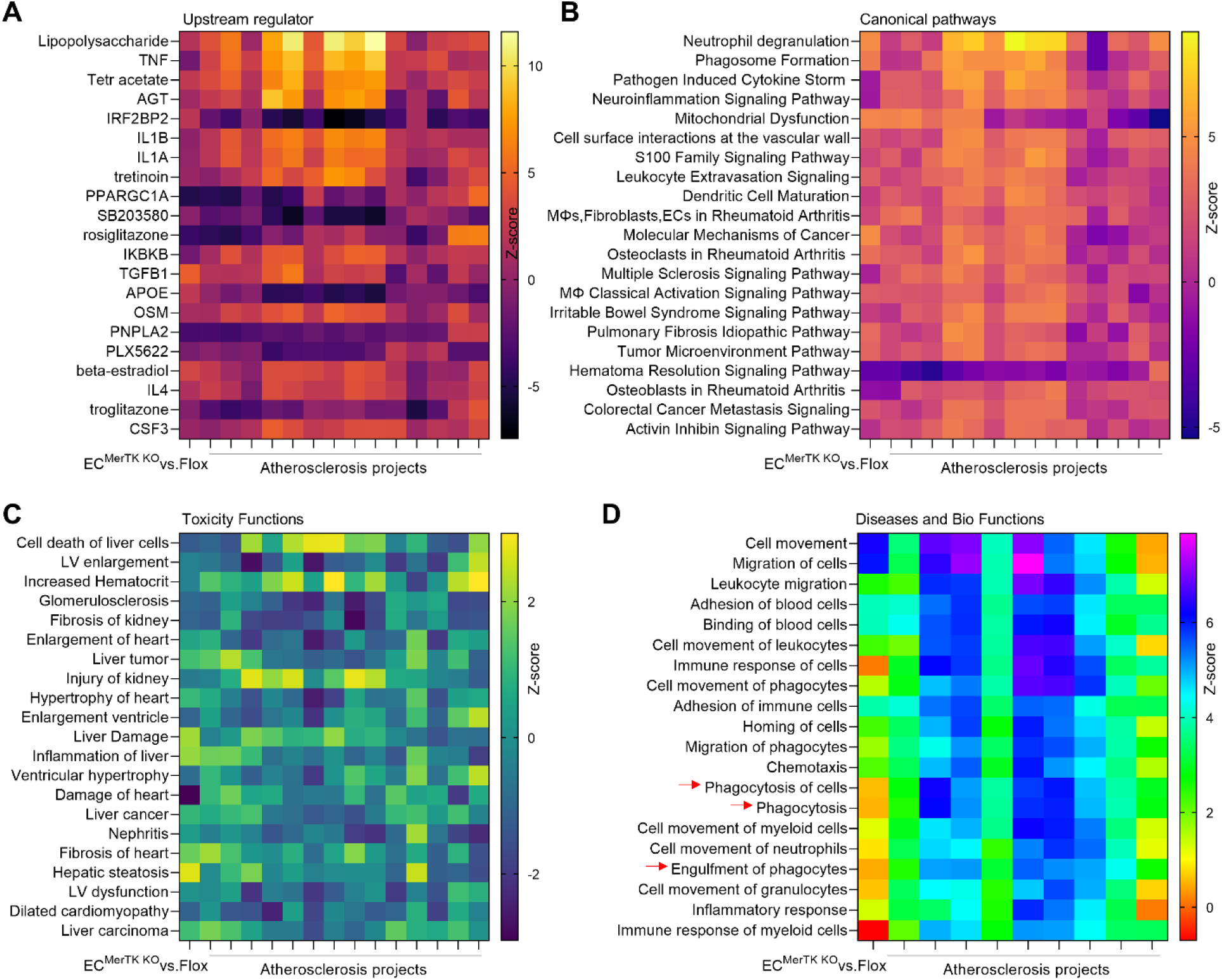
Multi-comparisons of proteomics (*MerTK^flox/flox^Tie2^Cre^* vs. *MerTK^flox/flox^*) with shared atherosclerosis projects. (**A**) Upstream regulators in *MerTK^flox/flox^Tie2^Cre^*vs. *MerTK^flox/flox^* compared with other atherosclerosis projects based on activation of z-score. (**B**) The volcano canonical pathways based on activation of z-score. (**C**) The toxicity functions analyses based on activation of z-score. (**D**) Diseases and biological functions analysis based on activation of z-score.

## Discussion

Atherosclerosis is a chronic disease characterized by lipid deposition and escalating inflammation in large arteries.^1–3^ A key feature of atherosclerotic lesions is the accumulation of apoptotic cells and subsequent formation of necrotic cell debris in the intimal space underneath the aortic EC monolayer. Emerging evidence indicates that aortic ECs have a robust capacity to perform efferocytosis, a process for the efficient clearance of apoptotic cells.^12–15,27,28^ MerTK, a critical efferocytosis receptor, is highly expressed in aortic ECs. ^12–15,27,28^ Although MerTK mutation in macrophages or MerTK global knockout models have been shown to exacerbate necrotic plaque formation^29^, the specific role of endothelial MerTK in atherosclerosis and its underlying mechanisms remain unclear. In this study, we combined big data analytics and microarray analyses of human atherosclerotic tissues with proteomics and a unique endothelial-specific MerTK knockout mouse model (*MerTK^flox/flox^Tie2^Cre^* mice) to elucidate this role. Our findings revealed that endothelial MerTK deficiency significantly accelerates the development of atherosclerosis. In addition, we identify signaling pathways of endothelial dysfunction, SMC phenotypic alterations, and mitochondrial dysfunction as the key drivers in endothelial MerTK-mediated atherosclerosis.

The accumulation of apoptotic cells and subsequent exacerbation of inflammation are the hallmarks of atherosclerosis.^1–3^ Apoptotic cells are primarily cleared through efferocytosis by professional phagocytes, including macrophages, neutrophils, dendritic cells, mast cells, and other monocytes.^8,9^ In contrast, non-professional phagocytes such as epithelial cells, fibroblasts, and ECs typically express fewer specialized receptors and demonstrate lower phagocytotic efficiency.^8,9^ However, recent studies have shown that aortic ECs express abundant MerTK, enabling them to efficiently perform efferocytosis, particularly under conditions with increased apoptotic cell burden. ^12–15,27,28^ ECs acts as a barrier between bloodstream and adjacent tissues, and endothelial dysfunction initiates various cardiovascular diseases, including atherosclerosis, aortic aneurysms and dissections, and vascular aging.^4–5^ Within atherosclerotic lesions, endothelial dysfunction is predominantly driven by necrotic plaque formation and proinflammatory responses.^1–5^ Such dysfunction is characterized by increased apoptotic cell accumulation, elevated oxidative stress, and sustained inflammatory response, all intricately linked with impaired efferocytosis. ^1–5,8,9^ Effective efferocytosis prevents secondary necrosis of apoptotic cells, thereby facilitating inflammation resolution, a process vital for restoring tissue integrity and maintaining physiological endothelial function.^8,9^ Oxidative stress generation, particularly mediated by NADPH oxidase, further contributes to vascular pathogenesis.^30^ NADPH oxidase is a multi-subunit enzyme complex comprising membrane bound components (p22^phox^ and gp91^phox^) and cytosolic subunits (p47^phox^, p67^phox^, and p40^phox^) that collectively catalyze the production of superoxide (O ^−•^) through electron transfer from NADPH.^31^ Our data showed that endothelial MerTK deficiency significantly enhances the expression of NADPH oxidase subunits (p22^phox^, p47^phox^, and gp91^phox^) and promotes the subsequent proinflammatory responses evident by elevated levels of IL-1β, MCP-1, NF-κB and IFN-γ. These findings indicate that endothelial MerTK impairment may represent a novel and independent pathogenic factor contributing to the progression of atherosclerosis.

NADPH oxidases and mitochondria are the two primary cellular sources of ROS production. NADPH oxidases are key generators of O_2_- and H_2_O_2_ while mitochondrion serves as another major source, producing mitochondrial ROS (Mito-ROS).^31^ Cytosolic H_2_O_2_ derived from mitochondria can phosphorylate and subsequently activate NADPH oxidases, amplifying cellular ROS production.^32–34^ This dynamic crosstalk between NADPH oxidase and mitochondrial ROS highlights an essential regulatory controlling intracellular redox homeostasis.^35,36^ Increasing evidence indicates that NADPH oxidase activation serves as a reliable marker for mitochondrial dysfunction, particularly when direct assessment of mitochondrial function in fixed tissues remains challenging. ^32–34^ Consequently, in our study of endothelial MerTK-mediated atherosclerosis, NADPH oxidase activation was utilized as an indirect indicator of mitochondrial function. In addition to NADPH oxidases and Mito-ROS, the MAPK signaling pathway significantly contributes to the regulation of oxidative stress and inflammatory responses during atherosclerosis development.^1–3^ ERK signaling regulates ROS and NO production, as well as activation of inflammatory gene transcription in atherosclerosis.^37^ Activation of P38 MAPK facilitates inflammatory responses, with its inhibition known to prevent the formation of atherosclerotic plaques.^39^ Likewise, JNK activation acerbates proinflammatory responses and promotes atherosclerotic plaque instability.^40^ Our findings demonstrate that endothelial MerTK deficiency activates both NADPH oxidases and MAPK family kinases (ERK, p38 MAPK and JNK), supporting the key role of endothelial MerTK in the development of atherosclerosis.

In summary, using big data analytics and microarray analysis of human atherosclerotic tissues, along with characterization of our unique *MerTK^flox/flox^Tie2^Cre^*genetic model of atherosclerosis and proteomic analysis, we have provided compelling evidence elucidating a novel mechanism involving endothelial MerTK in atherosclerosis development. Our findings highlight activated molecular pathways, including pro-inflammatory signaling, NADPH oxidase activation, and MAPK family pathways, as well as key cellular processes, such as endothelial dysfunction, SMC phenotypic alteration, and mitochondrial dysfunction, underlying endothelial MerTK-mediated atherosclerosis. Further study will investigate the therapeutic potential of restoring endothelial MerTK expression and function in the prevention and treatment of atherosclerosis.

## Data Availability

Data Sharing. The authors of this investigation declare that all the data, analytical methods, and study materials are available to the researchers. All the detailed information is available in the Supplemental Data.

## Acknowledgement

This work was supported by National Institutes of Health (NIH) grants R01HL162958 and the Georgia State University startup funding.

## Conflict of interest

The authors declare that they have no conflict of interest.

## Data Sharing

The authors of this investigation declare that all the data, analytical methods, and study materials are available to the researchers. All the detailed information is available in the Supplemental Data.

## Notes

### Competing Interest Statement

The authors have declared no competing interest.

### Clinical Trial

NA

### Author Declarations

A23050 at Georgia State University.

## Reference

1. Björkegren JL, Lusis AJ. Atherosclerosis: recent developments. Cell. 2022;185:1630–45.

2. Libby P. The changing landscape of atherosclerosis. Nature. 2021;592:524–33.

3. Gisterå A, Hansson GK. The immunology of atherosclerosis. Nature Reviews Nephrology. 2017;13:368–80.

4. Gimbrone Jr MA, García-Cardeña G. Endothelial cell dysfunction and the pathobiology of atherosclerosis. Circulation Research. 2016;118:620–36.

5. Howe KL, Fish JE. Transforming endothelial cells in atherosclerosis. Nature Metabolism. 2019;1:856–7.

6. Bennett MR, Sinha S, Owens GK. Vascular smooth muscle cells in atherosclerosis. Circulation Research. 2016;118:692–702.

7. Basatemur GL, Jørgensen HF, Clarke MC, Bennett MR, Mallat Z. Vascular smooth muscle cells in atherosclerosis. Nature Reviews Cardiology. 2019;16:727–44.

8. Doran AC, Yurdagul Jr A, Tabas I. Efferocytosis in health and disease. Nature Reviews Immunology. 2020;20:254–67.

9. Boada-Romero E, Martinez J, Heckmann BL, Green DR. The clearance of dead cells by efferocytosis. Nature Reviews Molecular Cell Biology. 2020;21:398–414.

10. De Couto G, Jaghatspanyan E, DeBerge M, Liu W, Luther K, Wang Y, Tang J, Thorp EB, Marbán E. Mechanism of enhanced MerTK-dependent macrophage efferocytosis by extracellular vesicles. Arteriosclerosis, Thrombosis, and Vascular Biology. 2019;39:2082–96.

11. Wang Y, Liu XY, Zhao WX, Li FD, Guo PR, Fan Q, Wu XF. NOX2 inhibition stabilizes vulnerable plaques by enhancing macrophage efferocytosis via MertK/PI3K/AKT pathway. Redox Biology. 2023;64:102763.

12. Liu S, Wu J, Stolarz A, Zhang H, Boerma M, Byrum SD, Rusch NJ, Ding Z. PCSK9 attenuates efferocytosis in endothelial cells and promotes vascular aging. Theranostics. 2023;13:2914–2929.

13. Wu J, Liu S, Banerjee O, Shi H, Xue B, Ding Z. Disturbed flow impairs MerTK-mediated efferocytosis in aortic endothelial cells during atherosclerosis. Theranostics. 2024;14:2427–2441.

14. Liu S, Wu J, Banerjee O, Xue B, Shi H, Ding Z. Big data analytics and scRNA-seq in human aortic aneurysms and dissections: role of endothelial MerTK. Theranostics. 2025;15:202–215.

15. Liu S, Wu J, Yang D, Xu J, Shi H, Xue B, Ding Z. Big data analytics for MerTK genomics reveals its double-edged sword functions in human diseases. Redox Biology.;70:103061.

16. 23.Searle BC, Pino LK, Egertson JD, Ting YS, Lawrence RT, MacLean BX. et al. Chromatogram libraries improve peptide detection and quantification by data independent acquisition mass spectrometry. Nat Commun. 2018;9:5128.

17. Graw S, Tang J, Zafar MK, Byrd AK, Bolden C, Peterson EC, Byrum SD. proteiNorm - A user-friendly tool for normalization and analysis of TMT and label-free protein quantification. ACS Omega. 2020;5:25625–33.

18. Döring Y, Manthey HD, Drechsler M, Lievens D, Megens RT, Soehnlein O, Busch M, Manca M, Koenen RR, Pelisek J, Daemen MJ, Lutgens E, Zenke M, Binder CJ, Weber C, Zernecke A. Auto-antigenic protein-DNA complexes stimulate plasmacytoid dendritic cells to promote atherosclerosis. Circulation. 2012;125:1673–83.

19. Gomez D, Baylis RA, Durgin BG, Newman AA, Alencar GF, Mahan S, St. Hilaire C, Müller W, Waisman A, Francis SE, Pinteaux E. Interleukin-1β has atheroprotective effects in advanced atherosclerotic lesions of mice. Nature Medicine. 2018;24:1418–29.

20. Georgakis MK, Van Der Laan SW, Asare Y, Mekke JM, Haitjema S, Schoneveld AH, De Jager SC, Nurmohamed NS, Kroon J, Stroes ES, De Kleijn DP. Monocyte-chemoattractant protein-1 levels in human atherosclerotic lesions associate with plaque vulnerability. Arteriosclerosis, Thrombosis, and Vascular Biology. 2021;41:2038–48.

21. Karunakaran D, Nguyen MA, Geoffrion M, Vreeken D, Lister Z, Cheng HS, Otte N, Essebier P, Wyatt H, Kandiah JW, Jung R. RIPK1 expression associates with inflammation in early atherosclerosis in humans and can be therapeutically silenced to reduce NF-κB activation and atherogenesis in mice. Circulation. 2021;143:163–77.

22. Poznyak AV, Grechko AV, Orekhova VA, Khotina V, Ivanova EA, Orekhov AN. NADPH oxidases and their role in atherosclerosis. Biomedicines. 2020;8:206.

23. Pejenaute Á, Cortés A, Marqués J, Montero L, Beloqui Ó, Fortuño A, Martí A, Orbe J, Zalba G. NADPH oxidase overactivity underlies telomere shortening in human atherosclerosis. International Journal of Molecular Sciences. 2020;21:1434.

24. Allahverdian S, Chaabane C, Boukais K, Francis GA, Bochaton-Piallat ML. Smooth muscle cell fate and plasticity in atherosclerosis. Cardiovascular Research. 2018;114:540–50.

25. Peng W, Cai G, Xia Y, Chen J, Wu P, Wang Z, Li G, Wei D. Mitochondrial dysfunction in atherosclerosis. DNA and Cell Biology. 2019;38:597–606.

26. Sanit J, Prompunt E, Adulyaritthikul P, Nokkaew N, Mongkolpathumrat P, Kongpol K, Kijtawornrat A, Petchdee S, Barrère-Lemaire S, Kumphune S. Combination of metformin and p38 MAPK inhibitor, SB203580, reduced myocardial ischemia/reperfusion injury in non-obese type 2 diabetic Goto-Kakizaki rats. Experimental and Therapeutic Medicine. 2019;18:1701–14.

27. Li Y, Wittchen ES, Monaghan-Benson E, Hahn C, Earp HS, Doerschuk CM, Burridge K. The role of endothelial MERTK during the inflammatory response in lungs. PLoS One. 2019;14:e0225051.

28. Happonen KE, Burrola PG, Lemke G. Regulation of brain endothelial cell physiology by the TAM receptor tyrosine kinase Mer. Communications Biology. 2023;6:916.

29. Cai B, Thorp EB, Doran AC, Sansbury BE, Daemen MJ, Dorweiler B, Spite M, Fredman G, Tabas I. MerTK receptor cleavage promotes plaque necrosis and defective resolution in atherosclerosis. The Journal of Clinical Investigation. 2017;127:564–8.

30. Schürmann C, Rezende F, Kruse C, Yasar Y, Löwe O, Fork C, Van De Sluis B, Bremer R, Weissmann N, Shah AM, Jo H. The NADPH oxidase Nox4 has anti-atherosclerotic functions. European Heart Journal. 2015;36:3447–56.

31. Zhang Y, Murugesan P, Huang K, Cai H. NADPH oxidases and oxidase crosstalk in cardiovascular diseases: novel therapeutic targets. Nature Reviews Cardiology. 2020;17:170–94.

32. Fukai T, Ushio-Fukai M. Cross-talk between NADPH oxidase and mitochondria: role in ROS signaling and angiogenesis. Cells. 2020;9:1849.

33. Daiber A, Di Lisa F, Oelze M, Kröller-Schön S, Steven S, Schulz E, Münzel T. Crosstalk of mitochondria with NADPH oxidase via reactive oxygen and nitrogen species signalling and its role for vascular function. British Journal of Pharmacology. 2017;174:1670–89.

34. Vendrov AE, Vendrov KC, Smith A, Yuan J, Sumida A, Robidoux J, Runge MS, Madamanchi NR. NOX4 NADPH oxidase-dependent mitochondrial oxidative stress in aging-associated cardiovascular disease. Antioxidants & Redox Signaling. 2015;23:1389–409.

35. Canugovi C, Stevenson MD, Vendrov AE, Hayami T, Robidoux J, Xiao H, Zhang YY, Eitzman DT, Runge MS, Madamanchi NR. Increased mitochondrial NADPH oxidase 4 (NOX4) expression in aging is a causative factor in aortic stiffening. Redox Biology. 2019;26:101288.

36. Du Z, Yang Q, Liu L, Li S, Zhao J, Hu J, Liu C, Qian D, Gao C. NADPH oxidase 2-dependent oxidative stress, mitochondrial damage and apoptosis in the ventral cochlear nucleus of D-galactose-induced aging rats. Neuroscience. 2015;286:281–92.

37. Wang C, Nie X, Zhang Y, Li T, Mao J, Liu X, Gu Y, Shi J, Xiao J, Wan C, Wu Q. Reactive oxygen species mediate nitric oxide production through ERK/JNK MAPK signaling in HAPI microglia after PFOS exposure. Toxicology and Applied Pharmacology. 2015;288:143–51.

38. Reustle A, Torzewski M. Role of p38 MAPK in atherosclerosis and aortic valve sclerosis. International Journal of Molecular Sciences. 2018;19:3761.

39. Elkhawad M, Rudd JH, Sarov-Blat L, Cai G, Wells R, Davies LC, Collier DJ, Marber MS, Choudhury RP, Fayad ZA, Tawakol A. Effects of p38 mitogen-activated protein kinase inhibition on vascular and systemic inflammation in patients with atherosclerosis. JACC: Cardiovascular Imaging. 2012;5:911–22.

40. Babaev VR, Yeung M, Erbay E, Ding L, Zhang Y, May JM, Fazio S, Hotamisligil GS, Linton MF. Jnk1 deficiency in hematopoietic cells suppresses macrophage apoptosis and increases atherosclerosis in low-density lipoprotein receptor null mice. Arteriosclerosis, Thrombosis, and Vascular Biology. 2016;36:1122–31.

